# Knowledge and Behaviors towards COVID-19 among University of Aleppo Students: An Online Cross-sectional Survey

**DOI:** 10.1101/2020.07.11.20151035

**Authors:** Aos Alhamid, Ziad Aljarad, Ahmad Alhamid

## Abstract

**Background:** Coronavirus Disease 2019 (COVID-19) is an infection caused by a novel coronavirus that affects respiratory tract. People’s awareness and knowledge of, and behavior and attitude toward COVID-19 are scarcely investigated, making medical literature related to this point poor. we aim to measure the knowledge of, and the reaction to COVID-19 among University of Aleppo students in Syria, and the determinants of their awareness and behavior regarding this disease.

**Materials and Methods:** This was an online, questionnaire-based cross-sectional study, that was conducted from 21st March to 30 March 2020. We included undergraduate students of the University of Aleppo (Syria). The questionnaire consisted of three sections: Demographics,knowledge and behaviours. Every participant’s knowledge was scored from 0-13 depending on the number of correct answers in the knowledge section. The correctness was judged depending on WHO recommendations at the time of questionnaire administration. P-value of 0.05 was considered significant.

**Results:** Among this well-educated and predominantly medical and health-related students, 682 (45.4%) students had a good knowledge level, which is somehow disappointing. The current study shows that 1st year students and non-medical specialties students and smokers had significantly lower knowledge levels than others. On contrary, residing with less people-which may indicate a higher socioeconomic status-, was associated with a higher knowledge level. We also found that commitment to preventive measures was in general satisfying and correlated significantly with knowledge level and gender in most cases.

**Conclusion:** Junior students, non-medical specialties, smokers and those who live with high number of people are vulnerable to less knowledge level and awareness campaigns should concentrate on them. Increasing awareness is useful to increase commitment to preventive measures, and groups that have less adherence to preventive measures, as described in detail, should be taken into consideration while designing public health responses. Finally, we should be aware of the negative impact of quarantine on public health to take it into consideration for current campaigns and future policies.

## Introduction

Coronavirus Disease 2019 (COVID-19) is an infection caused by a novel coronavirus that affects respiratory tract, it was first detected in Wuhan, China, in December 2019. Whole-genome sequencing of the virus declares that it is a betacoronavirus closely related to the Severe Acute Respiratory Syndrome (SARS) virus [1]. The origin of this virus is still questionable [2], but a zoonotic origin from bat and pangolin is suspected [3].

Most symptomatic infected people develop symptoms such as fever, fatigue, Dry cough, Anorexia, Myalgias, Sputum production and dyspnea. And 20% of patients may develop Acute respiratory distress syndrome (ARDS) [4]. Some studies have also reported gastrointestinal symptoms in 2–10% of cases such as diarrhea, abdominal pain, and vomiting [5,6] and neurological symptoms in 36.4 % of cases such as disturbed consciousness, headache, and paresthesia [7].

COVID-19 is transmitted from human-to-human through respiratory droplet, feco-oral route, and direct contact, and has an incubation period of 2-14 days [8].

COVID-19 is a rapidly expanding global threat, placing an intolerable burden on healthcare services. Number of cases is dramatically increasing every day. Until 21 March 2020, 292142 cases were detected worldwide, with 12784 deaths, and on 13 May 2020 4,337,562 cases were detected worldwide, with 292,450 deaths, 1,597,641 recovered cases, and 2,447,471 active cases [9]. It has been considered as a major threat to humans since December 31, 2019 [10], and lastly defined as a pandemic on 11 March 2020 [11].

To date, no vaccine or treatment has been approved for this disease, making preventive measures the most effective intervention [11]. This makes the public the main player in combating this pandemic.

People’s awareness and knowledge of, and behavior and attitude toward COVID-19 are scarcely investigated, making medical literature related to this point poor. in this study, we aim to measure the knowledge of, and the reaction to COVID-19 among University of Aleppo students, and the determinants of their awareness and behavior regarding this disease. This study is important to inform governments and concerned organizations about people thoughts and actions, especially the young, and may help to inform the upcoming public health measures and interventions.

## Materials and Methods

We conducted this cross-sectional study from 21st March to 30 March 2020. It was hard to make it a field-based study because of limited access to university students in this epidemic situation and because of the governmental measure to close the university temporally, so we made it a web-based study.

Therefore, we formed an online questionnaire and shared it with students on faculties Facebook, Telegram, and Whatsapp groups. The post introducing the questionnaire contained a background about the study, its aims, and some notes about filling the questionnaire, as well as the link to the questionnaire. The front page of the questionnaire contained a detailed informed consent statement for participation, and a button through it the participant can confirm the consent.

We included all students from the University of Aleppo, from both sexes and all ages who were studying in a bachelor degree or equivalent program, or in technical middle institutes. We excluded participants from other universities and graduated students. the questionnaire consisted of 3 parts: Demographics, awareness and knowledge of COVID-19, and behaviors toward COVID-19 and preventive measures. The Demographics part consisted of 12 questions about socio-economic and demographic characteristics, while the awareness and knowledge part included 13 questions on symptoms, transmission ways, preventive ways, therapy, and a question on the resource of participants information. Behavioral part contained 10 questions about the participants commitment to preventive measures and the impact of the quarantine on their daily habits.

Questions in knowledge part were true-false-I do not know questions. Participants who answered the question correctly took one point for each question, and who answered incorrectly or said “I don’t know “took zero points for the question. The correctness was considered depending on WHO recommendations on the day of questionnaire administration. [12] So, the degrees ranged from 0 to 13.

We conducted statistical analysis using SPSS (Version 22.0; SPSS Inc.: Chicago, IL, USA). Categorical variables were presented as frequencies and percentages, while continuous data was summarized as mean ± standard deviation (SD). We calculated Odds ratios (OR) with confidence intervals (CI) using logistic regression analysis. Whichever indicated, we used Chi-square, Fisher’s exact test and Student T-test tests to calculate the p-values. The p-value less than 0.05 was considered statistically significant.

This study was approved by the research committee of the University of Aleppo.

## Results

Among the 1502 students who responded, 834 (55.5%) were females, 881 (58.7%) studied a medical or health-related specialty, and 1423(94.7%) of them were single. Advanced students (who are in the 3rd year or more) constituted for the majority of the sample (904,60.2%). We found that 447 (29.8%) students were regular cigarettes or shisha smokers. More details on the demographic and socioeconomic characteristics of the participants are shown in table.1.

**Table 1:**
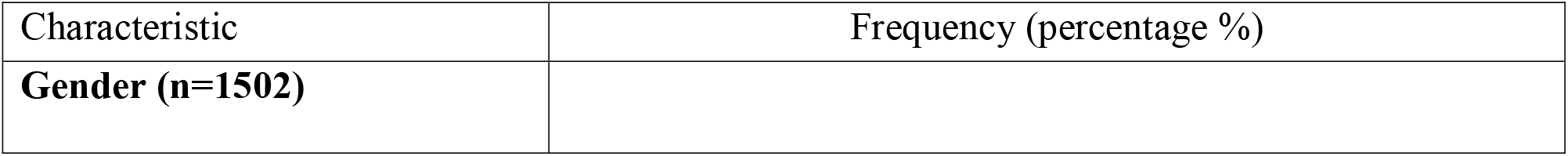
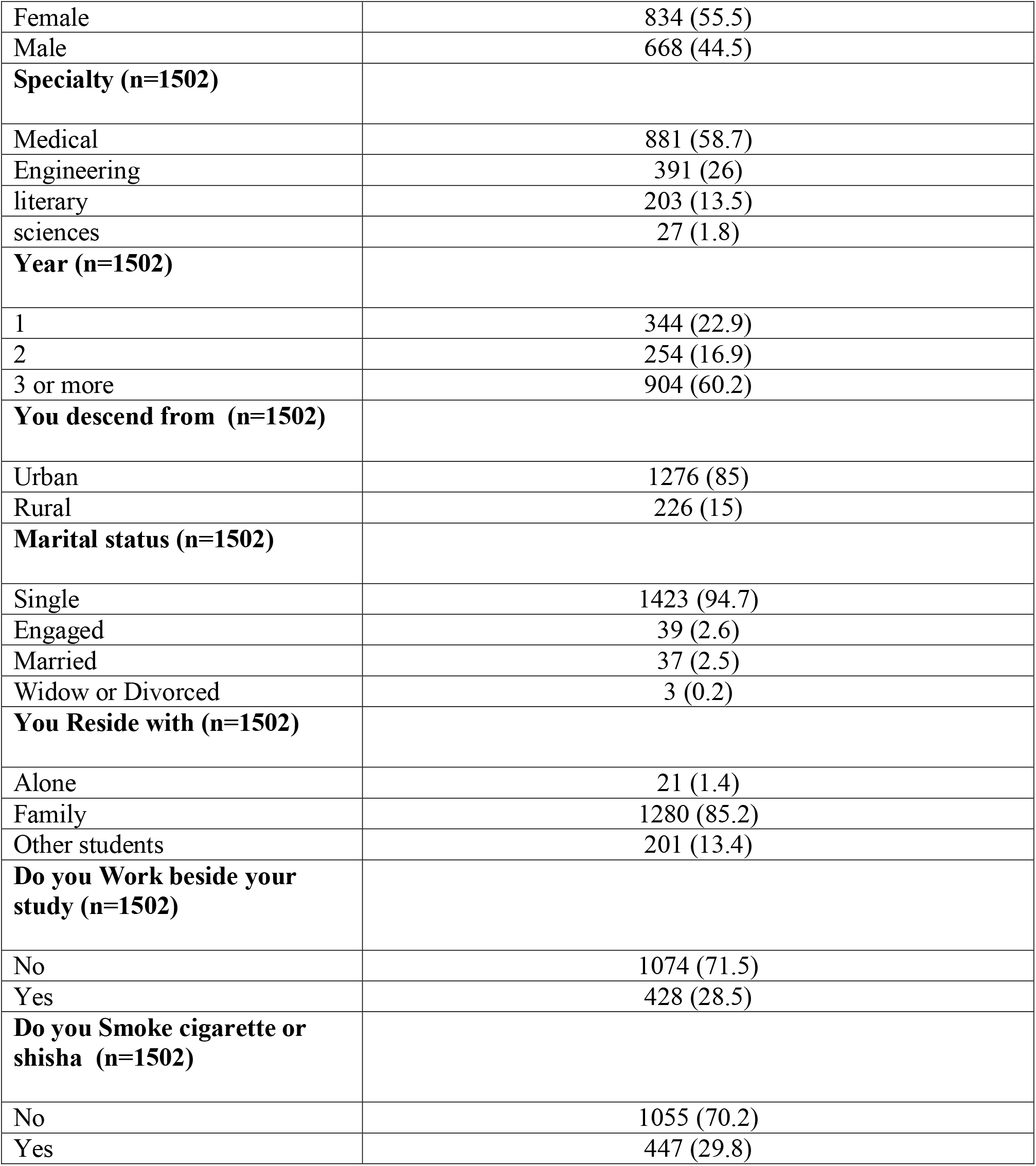
characteristics of participants and its frequencies. original

Table.2 demonstrates participants’ answers to the questions related to the knowledge and awareness of COVID-19. In most questions, the majority of respondents answered correctly to the questions, except for the question K9 (669, 44.6%) about the need to wear a mask outside, and question K13 (573,38.2%) which was about a rumor that spread virally through social media in Syria reporting a new easy diagnostic method for COVID-10 invented by an unknown Japanese scientist. We can also notice that 773 (51.6%) of participants answered correctly to question K7 about the possibility of being an asymptomatic COVID-19 patient, making this majority not vast and on borderline. The participants scores ranged from 0/13 to 13/13, with a mean score of COVID-19 knowledge of 9.2±1.8. 123 (8.2%) participants had a poor knowledge level, 697 (46.4%) had a moderate knowledge level, and 682 (45.4%) had a good knowledge level. Notably, social media was the main source of information about COVID-19 in 1041 (69.6%) of the study sample, while discussions with family and friends was the least common source of information (373, 23.9%).

**Table 2:**
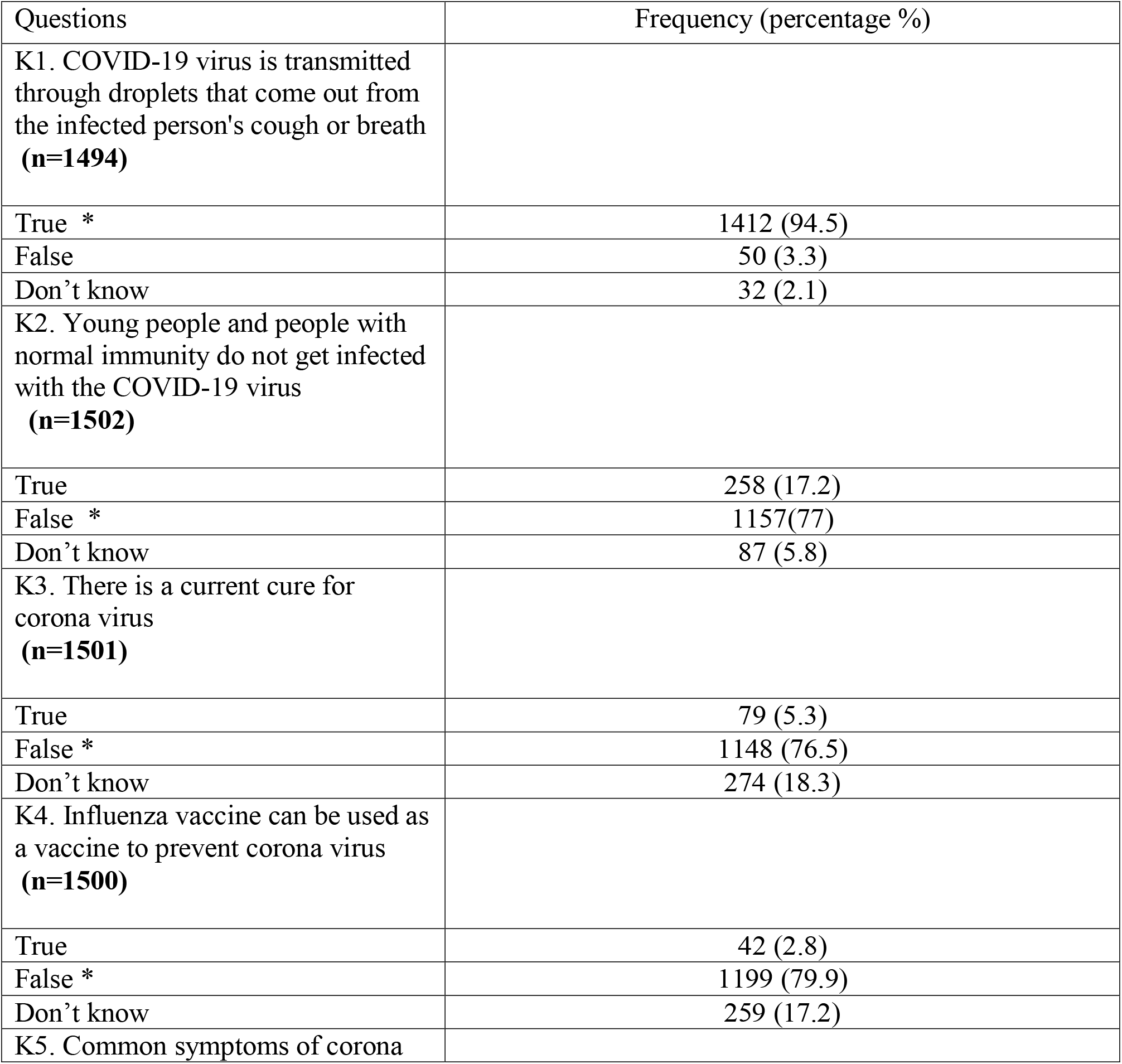
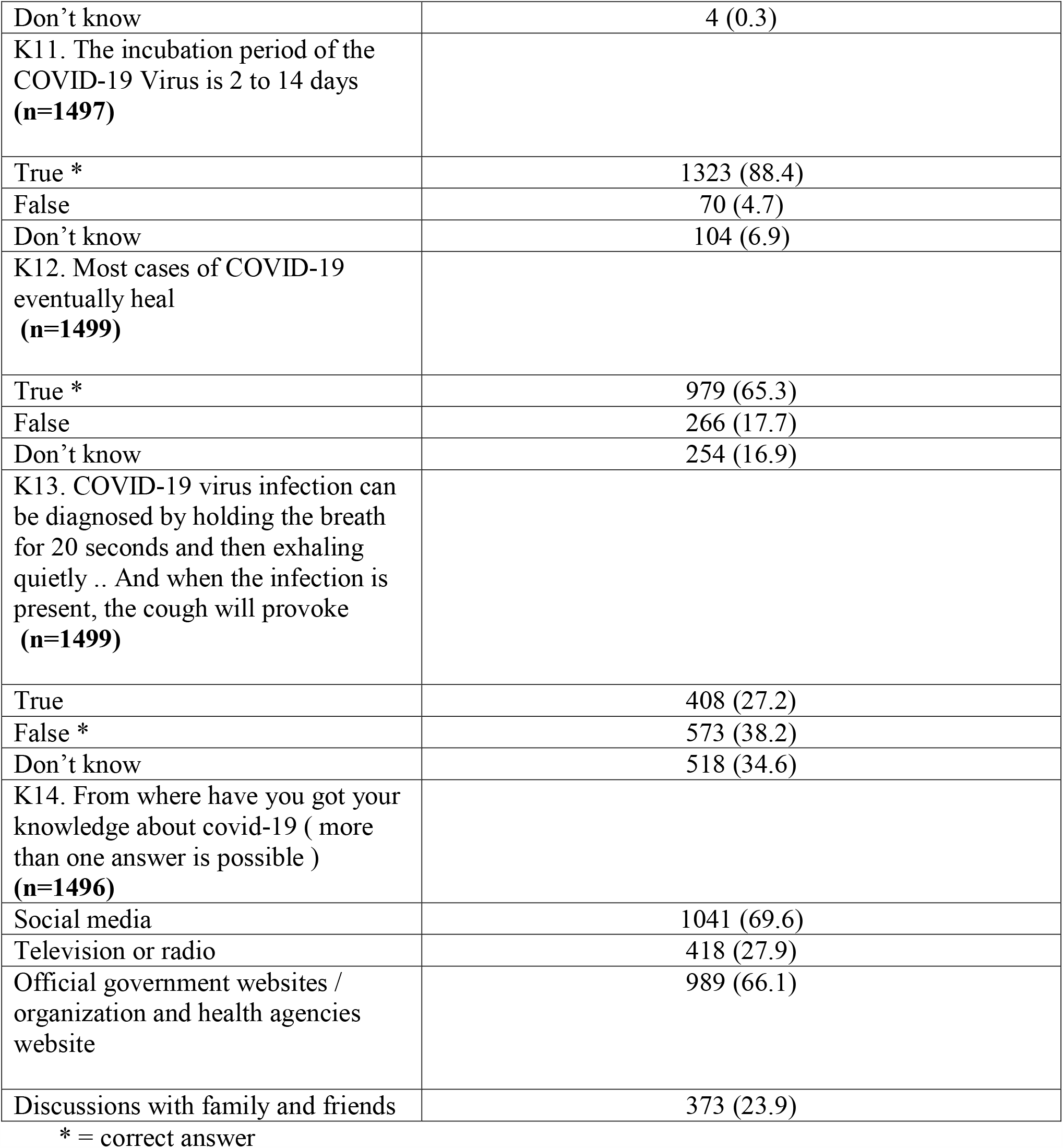
participants’ answers to the knowledge questions. original

Table.3 shows the behavior toward COVID-19 as reported by the participating students. We can notice that a considerable percentage of participants are committed to the preventive measures. 712 (47.7%) participants stopped going out to crowded places, 857 (57.2%) stopped going out with friends, and 896 (59.8%). In addition, as presented in table.3, there are some participants who were not used to do these behaviors in the first place, even before the pandemic. Most respondents had good hand-washing practices: 1380 (92.1%) students wash their hands more than 3 times a day, and 1323 (88.3%) students use soap in all their hand-washes. Only 109 (7.3%) always wear a mask when going out, while the immense majority did not (1120, 75.1%). 825 (55.3%) of participants sometimes keep a safe distance from others, while, unfortunately, 415 (27.8%) of them do not. Interestingly, 692 (46.1%) and 1323 (88.3%) of the study sample increased their consumption of food and internet, respectively, during the quarantine. It is also notable that 95 (21.3%) of smokers had higher smoking rate since the beginning of quarantine.

**Table 3:** participants’ answers to the behaviors questions. original

| Questions | Frequency (percentage %) |
| --- | --- |
| B1. Do you still go out to crowded places (such as markets, parks, and public squares)?<br>(n=1494) |  |
| Yes, as in the past | 18 (1.2) |
| Yes, less than in the past, but sometimes without necessity | 126 (8.4) |
| Yes, just for necessity | 477 (31.9) |
| I stopped going out to crowded places | 712 (47.7) |
| I primarily do not go out to crowded places | 161 (10.8) |
| B2. Do you still go out with your friends?<br>(n=1499) |  |
| Yes, as in the past | 46 (3.1) |
| Yes, less than in the past, but sometimes without necessity | 229 (15.2) |
| Yes, just for necessity | 306 (20.4) |
| I stopped going out with friends | 857 (57.2) |
| I primarily do not go out with friends | 61 (4.1) |
| B3. Do you still eat fast food?<br>(n=1499) |  |
| Yes, as in the past | 59 (3.9) |
| Yes, less than in the past, but sometimes without necessity | 123 (8.2) |
| Yes, just for necessity | 251 (16.7) |
| I stopped eating fast food | 896 (59.8) |
| I primarily do not eat fast food | 170 (11.3) |
| B4. How often do you wash your hands daily?<br>(n=1498) |  |
| never | 2 (0.1) |
| 1-3 | 116 (7.7) |
| More than 3 | 1380 (92.1) |
| B5. Do you always use soap when you wash your hands ?<br>(n=1498) |  |
| Always | 1323 (88.3) |
| Not always | 175 (11.7) |
| B6. Do you wear a mask when you go out on the street?<br>(n=1492) |  |
| Always | 109 (7.3) |
| Sometimes | 263 (17.6) |
| never | 1120 (75.1) |
| B7. Do you keep a safe distance of more than a meter when meeting anyone?<br>(n=1491) |  |
| Always | 251 (16.8) |
| Sometimes | 825 (55.3) |
| never | 415 (27.8) |
| B8. Has your consumption of food increased since the beginning of the home quarantine?<br>(n=1495) |  |
| Yes | 692 (46.1) |
| No | 803 (53.7) |
| B9. Has the number of hours spent on the Internet increased since the beginning of the home quarantine ?<br>(n=1499) |  |
| Yes | 1323 (88.3) |
| No | 176 (11.7) |
| B10. If you smoke a cigarette or shisha, have you increased your consumption of cigarettes or shisha |  |
| since the beginning of the home quarantine ?<br>(n=445) |  |
| Yes | 95 (21.3) |
| No | 350 (78.7) |

Table.4 shows the correlation between knowledge level and different variables. Specialty, year of study, who the student resides with, working status and smoking status were significantly associated with knowledge level.

**Table 4:** the correlation between variables and knowledge level. original

|  | Knowledge level |  |  |
| --- | --- | --- | --- |
|  | Poor<br>n (%) | Moderate<br>n (%) | Good<br>n (%) |
| **Gender** |  |  |  |
| female | 68 (55.3) | 407 (58.4) | 359 (52.6) |
| male | 55 (44.7) | 290 (41.6) | 323 (47.4) |
| **Specialty \*** |  |  |  |
| Medical | 29 (23.6) | 322 (46.2) | 530 (77.7) |
| Non-medical | 94 (76.4) | 375 (53.8) | 152 (22.3) |
| **Year \*** |  |  |  |
| 1 | 49 (39.8) | 204 (29.3) | 91 (13.3) |
| 2 | 22 (17.9) | 132 (18.9) | 100 (14.7) |
| 3 or more | 52 (42.3) | 361 (51.8) | 491 (72) |
| **Descent** |  |  |  |
| Rural | 14 (11.4) | 116 (16.6) | 96 (14.1) |
| Urban | 109 (88.6) | 581 (83.4) | 586 (85.9) |
| **Marital status** |  |  |  |
| Single | 120 (97.6) | 676 (97.0) | 666 (97.7) |
| Married and others | 3 (2.4) | 21 (3.0) | 16 (2.3) |
| **Reside with \*** |  |  |  |
| Alone | 1 (0.8) | 10 (1.4) | 10 (1.5) |
| Family | 108 (87.8) | 572 (82.1) | 600 (88) |
| With other friends | 14 (11.4) | 115 (16.5) | 72 (10.6) |
| **Working \*** |  |  |  |
| work | 45 (36.6) | 205 (29.4) | 178 (26.1) |
| Don't work | 78 (63.4) | 492 (70.6) | 504 (73.9) |
| **Smoking \*** |  |  |  |
| smoke | 49 (39.8) | 230 (33.0) | 168 (24.6) |
| Don't smoke | 74 (60.2) | 467 (67.0) | 514 (75.4) |
| **Number Resident partners** |  |  |  |
| 5 or less | 95 (77.2) | 573 (82.2) | 576 (84.5) |
| >5 | 28 (22.8) | 124 (17.8) | 106 (15.5) |
| **Age \*** |  |  |  |
| Mean | 20.6 | 20.9 | 21.2 |
| Std. Deviation | 2.1 | 2.2 | 1.9 |

\* P<0.05

table.5 reports the OR and 95% CI of the significantly related variables with knowledge level. Specialty, year of study, who you reside with, working, smoking and number of residency partners were significantly associated with knowledge level, depending on P-value or OR(95% CI).

**Table 5:** OR and 95% CI of the significantly related variables with knowledge level. original

| **Variable** | **OR (95% CI)** |
| --- | --- |
| **Moderate vs. poor knowledge level** |  |
| Specialty (medical vs. non-medical) | 2.783 (1.789 – 4.331) |
| Year of study (1<sup>st</sup> vs. 3<sup>rd</sup> or more) | 0.600 (0.392 – 0.918) |
| Year of study (2<sup>nd</sup> vs. 3<sup>rd</sup> or more) | 0.864 (0.505 – 1.478) |
| Who do you reside with (Alone vs. with other students) | 1.217 (0.145 – 10.236) |
| Who do you reside with (Family vs. with other students) | 0.645 (0.357 – 1.165) |
| Do you Work beside your study (no vs. yes) | 1.385 (0.927 – 2.069) |
| Do you smoke (no vs. yes) | 1.344 (0.907 – 1.994) |
| How many resident partners do you have ( 5 or less vs. more than 5 ) | 1.362 (0.856 – 2.166) |
| **Good vs. poor knowledge level** |  |
| Specialty (medical vs. non-medical) | 11.302 (7.180 – 17.791) |
| Year of study (1<sup>st</sup> vs. 3<sup>rd</sup> or more) | 0.197 (0.125 – 0.308) |
| Year of study (2<sup>nd</sup> vs. 3<sup>rd</sup> or more) | 0.481 (0.280 – 0.828) |
| Who do you reside with (Alone vs. with other students) | 1.944 (0.230 – 16.425) |
| Who do you reside with (Family vs. with other students) | 1.080 (0.588 – 1.984) |
| Do you Work beside your study (no vs. yes) | 1.634 (1.090 – 2.449) |
| Do you smoke (no vs. yes) | 2.026 (1.357 – 3.025) |
| How many resident partners do you have ( 5 or less vs. more than 5 ) | 1.602 (1.001 – 2.562) |

Table.6 demonstrates the association between each behavior and different variables, and table.7reports the OR and 95% CI of the significantly related variables with each behavior. Statistical significance was concluded evaluated by P-value or OR (95% CI). In general, gender, specialty and knowledge level were significant determinants of most behaviors. More details can be found in table.7.

**Table 6:** the correlation between variables and behaviors. original

|  | B1 |  | B2 |  | B3 |  |
| --- | --- | --- | --- | --- | --- | --- |
|  | Stopped<br>n (%) | have not<br>stopped<br>n (%) | Stopped<br>n (%) | have not<br>stopped<br>n (%) | Stopped<br>n (%) | have not<br>stopped<br>n (%) |
| **Gender** |  |  |  |  |  |  |
| female | 506 (71.1) | 237 (38.2) | 628 (73.3) | 169 (29.1) | 536 (59.8) | 216 (49.9) |
| male | 206 (28.9) | 384 (61.8)<br>* | 229 (26.7) | 412<br>(70.9)* | 360 (40.2) | 217<br>(50.1)* |
| **Specialty** |  |  |  |  |  |  |
| Medical | 422 (59.3) | 359 (57.8) | 522 (60.9) | 319 (54.9) | 519 (57.9) | 265 (61.2) |
| Non-medical | 290 (40.7) | 262 (42.2) | 335 (39.1) | 262<br>(45.1)* | 377 (42.1) | 168 (38.8) |
| **Year** |  |  |  |  |  |  |
| 1 | 161 (22.6) | 131 (21.1) | 202 (23.6) | 125 (21.5) | 198 (22.1) | 88 (20.3) |
| 2 | 121 (17.0) | 100 (16.1) | 140 (16.3) | 103 (17.7) | 143 (16.0) | 89 (20.6) |
| 3 or more | 430 (60.4) | 390 (62.8) | 515 (60.1) | 353 (60.8) | 555 (61.9) | 256 (59.1) |
| **Descent** |  |  |  |  |  |  |
| Rural | 97 (13.6) | 99 (15.9) | 108 (12.6) | 105 (18.1) | 138 (15.4) | 52 (12.0) |
| Urban | 615 (86.4) | 522 (84.1) | 749 (87.4) | 476<br>(81.9)* | 758 (84.6) | 381 (88.0) |
| **Marital status** |  |  |  |  |  |  |
| Single | 696 (97.8) | 602 (96.9) | 839 (97.9) | 562 (96.7) | 876 (97.8) | 416 (96.1) |
| Married and<br>others | 16 (2.2) | 19 (3.1) | 18 (2.1) | 19 (3.3) | 20 (2.2) | 17 (3.9) |
| **Reside with** |  |  |  |  |  |  |
| Alone | 11 (1.5) | 9 (1.4) | 13 (1.5) | 8(1.4) | 16 (1.8) | 4 (0.9) |
| Family | 614 (86.2) | 525 (84.5) | 750 (87.5) | 471 (81.1) | 763 (85.2) | 371 (85.7) |
| With other<br>friends | 87 (12.2) | 87 (14.0) | 94 (11.0) | 102<br>(17.6)* | 117 (13.1) | 58 (13.4) |
| **Working** |  |  |  |  |  |  |
| work | 163 (22.9) | 234 (37.7) | 198 (23.1) | 214 (36.8) | 254 (28.3) | 124 (28.6) |
| Don't work | 549 (77.1) | 387<br>(62.3)* | 659 (76.9) | 367<br>(63.2)* | 642 (71.7) | 309 (71.4) |
| **Smoking** |  |  |  |  |  |  |
| smoke | 164 (23.0) | 244 (39.3) | 164 (19.1) | 275 (47.3) | 248 (27.7) | 165 (38.1) |
| Don't smoke | 548 (77.0) | 377 (60.7)* | 693 (80.9) | 306 (52.7)* | 684 (72.3) | 268 (61.9)* |
| **Number of Resident partners** |  |  |  |  |  |  |
| 5 or less | 580 (81.5) | 524 (84.4) | 704 (82.1) | 489 (84.2) | 753 (84.0) | 359 (82.9) |
| >5 | 132 (18.5) | 97 (15.6) | 153 (17.9) | 92 (15.8) | 143 (16.0) | 74 (17.1) |
| **Age** |  |  |  |  |  |  |
| Mean | 20.9 | 21.2 | 20.8 | 21.3 | 21.0 | 21.2 |
| Std. Deviation | 2.0 | 2.1 * | 1.9 | 2.2 * | 2.0 | 2.3 |
| **Knowledge level** |  |  |  |  |  |  |
| Poor | 46 (6.5) | 58 (9.3) | 68 (7.9) | 45 (7.7) | 74 (8.3) | 31 (7.2) |
| Moderate | 332 (46.6) | 299 (48.1) | 387 (45.2) | 285 (49.1) | 406 (45.3) | 219 (50.6) |
| good | 334 (46.9) | 264 (42.5) | 402 (46.9) | 251 (43.2) | 416 (46.4) | 183 (42.3) |

\* P<0.05

**Table 7:** OR and 95% CI of the significantly related variables with behaviors. original

| Variable | OR (95% CI) |
| --- | --- |
| **B1 ( stopped vs. have not stopped )** |  |
| Gender ( female vs. male ) | 3.980 (3.165 – 5.004) |
| Do you Work beside your study (yes vs. no) | 0.491 (0.387 – 0.623) |
| Do you smoke (yes vs. no) | 0.462 (0.365 – 0.586) |
| Knowledge level (moderate vs. poor) | 1.400 (0.922 – 2.125) |
| Knowledge level (good vs. poor) | 1.595 (1.049 – 2.426) |
| **B2 ( stopped vs. have not stopped )** |  |
| Gender ( female vs. male ) | 6.686 (5.289 – 8.452) |
| Specialty (medical vs. non-medical) | 1.280 (1.034 – 1.584) |
| You descend from ( urban vs. rural ) | 1.530 (1.142 – 2.049) |
| Who do you reside with (Alone vs. with other students) | 1.763 (0.700 – 4.443) |
| Who do you reside with (Family vs. with other students) | 1.728 (1.276 – 2.339) |
| Do you Work beside your study (yes vs. no) | 0.515 (0.409 – 0.650) |
| Do you smoke (yes vs. no) | 0.263 (0.208 – 0.333) |
| **B3 ( stopped vs. have not stopped )** |  |
| Gender ( female vs. male ) | 1.496 (1.187 – 1.884) |
| Do you smoke (yes vs. no) | 0.622 (0.488 – 0.793) |
| **B4 ( more than 3 vs. 3 or less )** |  |
| Gender ( female vs. male ) | 2.737 (1.838 – 4.075) |
| Year of study (1<sup>st</sup> vs. 3<sup>rd</sup> or more) | 0.589 (0.379 – 0.915) |
| Year of study (2<sup>nd</sup> vs. 3<sup>rd</sup> or more) | 0.591 (0.363 – 0.961) |
| Do you smoke (yes vs. no) | 0.591 (0.402 – (0.869) |
| Knowledge level (moderate vs. poor) | 2.114 (1.221 – 3.658) |
| Knowledge level (good vs. poor) | 3.228 (1.811 – 5.754) |
| **B5 (No vs. Yes)** |  |
| Gender ( female vs. male ) | 0.382 (0.275 – 0.531) |
| Specialty (medical vs. non-medical) | 0.646 (0.471 – 0.886) |
| You descend from ( urban vs. rural ) | 0.597 (0.404 – 0.883) |
| Do you Work beside your study (yes vs. no) | 1.620 (1.166 – 2.250) |
| Do you smoke (yes vs. no) | 1.503 (1.082 – 2.086) |
| Knowledge level (moderate vs. poor) | 0.535 (0.329 – 0.872) |
| Knowledge level (good vs. poor) | 0.363 (0.219 – 0.603) |
| **B6 (always vs. never)** |  |
| Gender ( female vs. male ) | 1.819 (1.197 – 2.766) |
| Specialty (medical vs. non-medical) | 0.445 (0.299 – 0.663) |
| Do you Work beside your study (yes vs. no) | 0.796 (0.520 – 1.219) |
| Knowledge level (moderate vs. poor) | 2.682 (1.352 – 5.319) |
| Knowledge level (good vs. poor) | 2.241 (1.441 – 3.485) |
| **B6 (sometimes vs. never)** |  |
| Gender ( female vs. male ) | 1.142 (0.871 – 1.498) |
| Specialty (medical vs. non-medical) | 0.633 (0.483 – 0.829) |
| Do you Work beside your study (yes vs. no) | 0.638 (0.480 – 0.848) |
| Knowledge level (moderate vs. poor) | 1.550 (0.931 – 2.579) |
| Knowledge level (good vs. poor) | 1.624 (1.221 – 2.159) |
| **B7 (always vs. never)** |  |
| Gender ( female vs. male ) | 2.092 (1.517 – 2.883) |
| You descend from ( urban vs. rural ) | 0.782 (0.514 – 1.191) |
| Do you smoke (yes vs. no) | 1.064 (0.764 – 1.483) |
| How many resident partners do you have ( 5 or less vs. more than 5 ) | 1.199 (0.806 – 1.784) |
| Knowledge level (moderate vs. poor) | 0.624 (0.351 – 1.111) |
| Knowledge level (good vs. poor) | 1.139 (0.819 – 1.584) |
| Year of study (1<sup>st</sup> vs. 3<sup>rd</sup> or more) | 0.771 (0.525 – 1.133) |
| Year of study (2<sup>nd</sup> vs. 3<sup>rd</sup> or more) | 1.116 (0.736 – 1.693) |
| **B7 (sometimes vs. never)** |  |
| Gender ( female vs. male ) | 1.730 (1.364 – 2.195) |
| You descend from ( urban vs. rural ) | 0.634 (0.461 – 0.871) |
| Do you smoke (yes vs. no) | 1.446 (1.121 – 1.865) |
| How many resident partners do you have ( 5 or less vs. more than 5 ) | 1.536 (1.134 – 2.081) |
| Knowledge level (moderate vs. poor) | 0.481 (0.314 – 0.738) |
| Knowledge level (good vs. poor) | 1.092 (0.851 – 1.401) |
| Year of study (1<sup>st</sup> vs. 3<sup>rd</sup> or more) | 0.705 (0.531 – 0.937) |
| Year of study (2<sup>nd</sup> vs. 3<sup>rd</sup> or more) | 0.843 (0.608 – 1.168) |
| **B8 ( yes vs. no )** |  |
| Gender ( female vs. male ) | 1.574 (1.280 – 1.935) |
| Marital status (single vs. married and other ) | 2.047 (1.033 – 4.058) |
| Who do you reside with (Alone vs. with other students) | 1.148 (0.463 – 2.846) |
| Who do you reside with (Family vs. with other students) | 0.703 (0.521 – 0.947) |
| **B9 ( yes vs. no )** |  |
| Gender ( female vs. male ) | 1.510 (1.101 – 2.070) |
| You descend from ( urban vs. rural ) | 1.662 (1.125 – 2.457) |
| Marital status (single vs. married and other ) | 3.384 (1.688 – 6.784) |
| Knowledge level (moderate vs. poor) | 1.375 (0.810 – 2.334) |
| Knowledge level (good vs. poor) | 1.736 (1.012 – 2.980) |
| **B10 ( yes vs. no )** |  |
| Gender ( female vs. male ) | 0.541 (0.322 – 0.907) |
| Knowledge level (moderate vs. poor) | 0.374 (0.195 – 0.720) |
| Knowledge level (good vs. poor) | 0.292 (0.145 – 0.588) |

## Discussion

To the best of our knowledge, this is the first study to measure awareness of and behaviors and attitudes toward COVID-19 pandemic in Syria. Among this well-educated and predominantly medical and health-related students, 682 (45.4%) students had a good knowledge level, which is somehow disappointing. The current study shows that 1^st^ year students and non-medical specialties students and smokers had lower knowledge levels than others, which strongly suggests to concentrate on these groups in awareness campaigns. The lower knowledge level among the first mentioned two groups may be explained by the less medical education they received compared to advanced or medical students. The lower knowledge level among smokers seems to be an association with lower health awareness or less interest in personal and public health. On contrary, residing with less people-which may indicate a higher socioeconomic status-, was associated with a higher knowledge level.

Regarding adherence to preventive measures, we can notice that always females higher commitment levels than males. Knowledge level was associated with behavior in most but not all aspects, and behavior was also correlated with the same factors that were found correlated with knowledge, like scholar year, specialty, work, urban or rural descent, smoking and who and how many you reside with. This indicates that higher awareness leads in general to better action.

The quarantine has increased the consumption of food, internet in 692 (46.1%), 1323 (88.3%) of participants respectively, and increased tobacco consumption in 95 (21.3%) of smokers, which means that the quarantine may lead to future side effects on public health.

Unfortunately, to our knowledge, there is no peer-reviewed reports on the same topic of our study published from neighboring countries. However, we found a similar Chinese study performed on public, that reported a high knowledge level with a correct answer rates of 70.2-98.6%. [13], This might be because China was the first country to be affected with COVID-19, and much more than Syria.

Our study has major limitations. First, the questionnaire is delivered online, which makes quality control challenging, and reduces the accessibility of some vulnerable groups less probable. In addition, our tools were not well developed and standardized because of limited time and accessibility to the target population.

## Conclusion

In summary, our study reports a somehow low knowledge level of COVID-19 if we took into consideration that the target population was university students. Junior students, non-medical specialties, smokers and those who live with high number of people are vulnerable to less knowledge level and awareness campaigns should concentrate on them. Increasing awareness is useful to increase commitment to preventive measures, and groups that have less adherence to preventive measures, as described in detail, should be taken into consideration while designing public health responses. Finally, we should be aware of the negative impact of quarantine on public health to take it into consideration for current campaigns and future policies.

## Data Availability

Data are available upon request by contacting the corresponding author

## Data Availability

Data are available upon request by contacting the corresponding author.

